# Atopic dermatitis and risk of 14 site-specific cancers: a Mendelian randomization study

**DOI:** 10.1101/2023.03.30.23287958

**Authors:** Qiang Liu, Li Chen, Yipeng Wang, Xiangyu Wang, Sarah J Lewis, Jing Wang

**Author notes:** Corresponding Authors **Jing Wang**, Department of Breast Surgical Oncology, National Cancer Center/National Clinical Research Center for Cancer/Cancer Hospital, Chinese Academy of Medical Sciences and Peking Union Medical College, Beijing, 100021, China., **Sarah J Lewis**, MRC Integrative Epidemiology Unit, Population Health Sciences, Bristol Medical School, University of Bristol, Bristol, UK; Bristol Dental School, University of Bristol, Bristol, UK. These authors contributed equally to this work (Sarah J Lewis, Jing Wang).

## Abstract

**IMPORTANCE:** Atopic dermatitis (AD) accounts for a large proportion of the burden of skin disease with a prevalence of around 10% among adults worldwide. In addition, systematic reviews and meta-analyses have found that AD is associated with cancer risk at several sites, if found to be causal this could highlight potential treatment targets to reduce cancer risk.

**OBJECTIVE:** To assess the potential causative link between AD and 14 site-specific cancers in a two-sample Mendelian randomization study.

**EXPOSURE:** Atopic dermatitis

**DESIGN, SETTING, AND PARTICIPANTS:** From the largest genome-wide association study (GWAS) of AD (10,788 cases and 30,047 non-cases), genetic variants highly associated (P < 5E-08) with AD in European population were selected as instrumental variables (IVs). Data from large cancer consortia, as well as the UK Biobank study(n=442,239) and the FinnGen study (n=218,792) were employed to assess genetic associations with 14 site-specific cancers and overall cancer. A set of complementary approaches and sensitivity analyses were carried out to examine the robustness of our results. In addition, associations for the same cancer site from different data sources were combined using meta-analyses.

**RESULTS:** We discovered no strong causal evidence of AD on the risk of overall cancer, with effect estimates close to zero. After Benjamini–Hochberg correction, the inverse weighted method indicated no association of AD on overall cancer risk in both the UK biobank (OR, 1.00; 95%CI, 0.94-1.06; FDR, 0.98) and FinnGen studies (OR, 0.96; 95%CI, 0.92, 1.02; FDR, 0.68). No strong evidence of association was found between genetically predicted AD and the risk of any other site-specific cancers.

**CONCLUSIONS AND RELEVANCE:** Our MR investigation does not support a causal effect of AD on cancer risk. This finding has important implications for the prevention and management of both AD and cancer, as it reduces the concern of potential adverse effects of AD on cancer outcomes.

## Introduction

Atopic dermatitis (AD), commonly known as atopic eczema, is a skin condition that is chronic, inflammatory, and eczematous. The name eczema relates to the disease’s eczematous characteristics, which are long lasting; serous leaking, and blistering in addition to erythema and scaling^1^. The prevalence of AD has increased in many countries and has a significant impact on the health of patients and their families^1-3^, affecting up to 20% of children and up to 10% of adults^4^. Cancer is one of the leading causes of mortality in the world, accounting for over 10 million fatalities in 2020, or roughly one out of every six deaths^5^. The chronic inflammation, which occurs with eczema, can damage DNA and potentially alter the risk of mutations that lead to cancer. Furthermore, some treatments for AD may suppress the immune system and thereby altering the risk of cancer. Previous epidemiological research has provided interesting findings on the association between AD and cancer risk. A systematic review and meta-analysis found observational evidence of associations between AD and increased risk of keratinocyte carcinoma and kidney cancer and lower risk of lung and respiratory system cancers^6^, while another meta-analysis found that AD was inversely associated with brain cancer^7^. Recent cohort studies conducted in England and Denmark revealed no indication of an association between AD and most cancers, with the exception of lymphoma^8^. So far the evidence connecting AD and cancer risk in humans has been based on observational studies, which are susceptible to confounding and reverse causation. As a result, the causal role of AD in the development of cancer remains uncertain.

Mendelian randomization (MR) is an effective epidemiological method that uses genetic variants as a tool to improve causal inference^9^. Because genetic variations are randomly assigned at conception and hence not influenced by environmental and self-adapted variables, MR is naturally not prone to confounding. Furthermore, because germline genotypes are unaffected by biological perturbations which occur due to illness, this strategy can avoid the problem of reverse causality. However, MR analyses of AD with the risk of site-specific cancers have not been yet been conducted. Herein, we conducted an MR analysis to assess whether there are causal relations between AD with risk of overall cancer and 14 site-specific cancers.

## Methods Data sources

**Fig. 1** depicts a summary of the study design. We used summary-level data for AD and cancers from the UK biobank, FinnGen study, and several international consortia to conduct two-sample MR. These consortia included the Breast Cancer Association Consortium^10^ (122,977 cases and 105,974 controls), the Ovarian Cancer Association Consortium^11^ (25,509 cases and 40,941 controls), International Lung Cancer Consortium^12^ (11,348 cases and 15,861 controls), and the Prostate Cancer Association Group to Investigate Cancer Associated Alterations in the Genome consortium (79,148 cases and 61,106 controls)^13^. Summary information from the largest European population GWAS for AD conducted to date was used for the SNP-exposure estimates. Genome-wide association studies (GWAS) of AD, pan-cancer and site-specific cancers in UK Biobank and FinnGen studies were accessed through the MRC Integrative Epidemiology Unit (IEU) Open GWAS database^14-16^ (https://gwas.mrcieu.ac.uk/). GWAS summary level data for our positive control outcome-asthma and our negative control outcome body height were also accessed through the MRC IEU pen GWAS database. **Table. S1** details Information of all included studies and international consortiums involved in this study. **Table. S2** presents sources and definitions of site-specific cancers. This research was approved by the institutional review board at the Cancer Hospital, Chinese Academy of Medical Sciences and Peking Union Medical College, and written informed consent was waived because it utilised data from publicly available, de-identified datasets.

**Figure 1.**
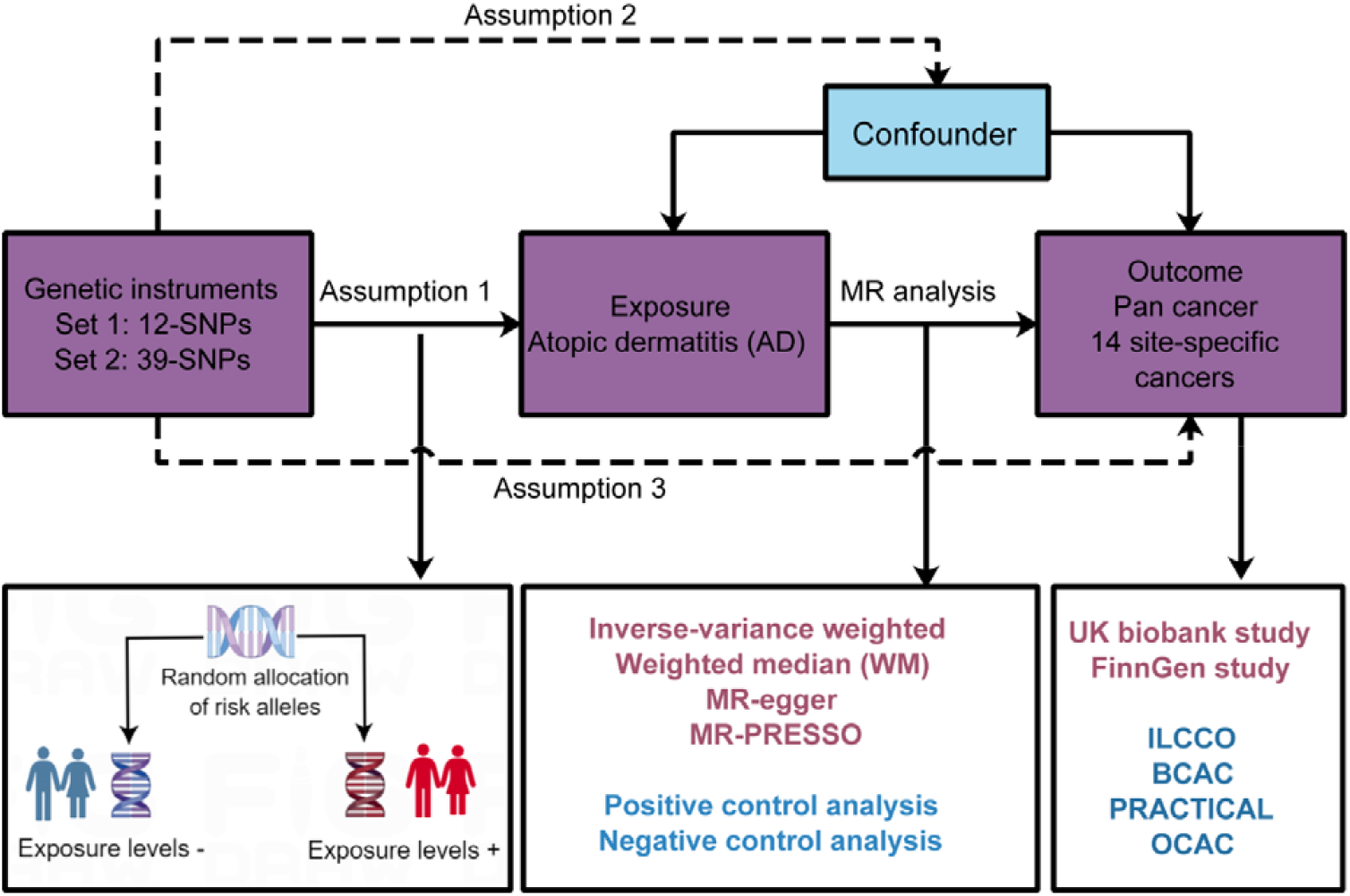
Overview of the study design.

### Genetic instrument selection

The genome-wide summary statistics for AD used for this study contains 10,788 cases and 30,047 controls from 20 studies of European ancestry. Two strategies were employed in the process of deciding upon the instruments to utilise. Firstly, SNPs associated with AD at the genome-wide significance level (P < 5 × 10^−8^) were selected (Set 1-IVs). Secondly, we used genetic instruments based a lower significance threshold (P < 5*10^−6^) in order to increase the explained phenotypic variance and therefore the statistical power (Set 2-IVs). When specific single-nucleotide polymorphisms (SNPs) were not included in the outcome datasets, an appropriate proxy in strong linkage disequilibrium r^2^=0.8 was employed as a replacement. After excluding SNPs which were in linkage disequilibrium (r^2^ ≥ 0.001) with another SNP in our instrument, 12 SNPs (Set 1-IVs) and 39 SNPs (Set 2-IVs), with F statistics more than 10 were selected as instrumental variables (IVs) (**Table. S3**). The combined F statistic for the IVs was calculated using the formula provided by Burgess et al.^17^, yielding values of 48.6 and 37.4 for Set 1-IVs and Set 2-IVs, respectively. We calculated the proportion of variance in the exposure explained by the genetic variants (R^2^) by adding R^2^ of each SNP together. The instrumental variable accounted for 1.4% (Set 1) and 3.5% (Set 2) of the variance for AD, respectively. In the primary analysis, we scaled the odds ratios (ORs) and 95% confidence intervals (CIs) of the relationships to a 1-unit increase in the log-transformed odds of genetic liability to AD.

### Statistical analysis

We estimated odds ratios (OR) and 95% CI of the effect of AD on malignancies using SNP-exposure and SNP-outcome beta coefficients and SEs. By dividing the SNP-outcome relationship by the SNP-exposure association, we were able to get the causal estimate for individual SNPs (Wald ratio). The causal estimate for multi-SNP instruments was computed using inverse-variance weighted (IVW)-MR, which takes an average of Wald ratios across SNPs, weighted by the SNP-exposure beta coefficients ^18^. IVW-MR implies that all instruments are genuine or that pleiotropy is balanced, and we assumed linear associations between SNPs and exposure and between SNPs and outcome ^19^. The core assumptions of MR^9,20,21^, which can be tested via sensitivity analyses, are that the instrument: predicts exposure; is unaffected by confounders of the exposure/outcome relationship; and affects the outcome only through exposure, meaning is there is no horizontal pleiotropy. We performed sensitivity analyses to test the robustness of our findings and the possibility of the assumptions being violated, most notably horizontal pleiotropy. We estimated Cochran’s Q-statistic for effect heterogeneity between SNPs. To identify any horizontal pleiotropy, the intercept test of MR-Egger regression was performed. We used complementary approaches which make different assumptions relating to pleiotropy to calculate MR estimates as sensitivity analyses, including weighted-median MR ^22^, MR-Egger^23^, although prone to imprecision; and simple mode method (estimates based on the cluster with the largest number of SNPs) ^24^. To detect outlier SNPs that may have distorted our findings, we examined per-SNP causal estimates (scatter, forest plots) and conducted leave-one-out analyses. We used MR Pleiotropy Residual Sum and Outlier (MR-PRESSO) method to detect outlying SNPs with global-pleiotropy and SNP-outlier tests p<0.05^25^. Using the PhenoScanner database^26^, we investigated whether SNPs were associated with other relevant traits (possible confounders, smoking, alcohol intake) and we excluded potentially pleiotropic SNPs based on this.^17^ Positive and negative control outcome analyses were employed to assess the potential bias due to horizontal pleiotropy and selection bias^20^. To correct for multiple testing, the Benjamini-Hochberg method was used, which controls the false discovery rate (FDR). R software was used for data preparation and analysis (R version 4.2.2). MR analyses were carried out with the TwoSampleMR package^27^. The mRnd Mendelian randomization power calculation online tool^28^ was used to determine statistical power. Meta-analysis was used to combine MR estimates from different data sources for the same cancer site. This study follows the guidelines for Strengthening the Reporting of Observational Studies in Epidemiology (STROBE) guideline (**Supplement Checklist**).

## Results

In analyses using consortia data (breast cancer, prostate cancer, ovarian cancer, endometrial cancer) and overall cancer in UK biobank, we had more than 90% power at a significance level of 0.05 to detect an OR of 1.25, whereas relatively large magnitudes of associations were necessary to detect strong evidence of effect for some site-specific cancer analyses, this is presented in **Table. S4**). We found no strong association of AD on overall cancer risk in both the UK biobank (OR, 1.00; 95%CI, 0.94-1.06; FDR, 0.98) and FinnGen studies (OR, 0.96; 95%CI, 0.92, 1.02; FDR, 0.68). **Fig. 2 and Fig. S1** show the relationships of genetically predicted AD 14 site-specific cancers in the UK biobank and large international consortia using two sets of IVs. After Benjamini-Hochberg adjustment, genetically predicted AD was not related with the 14 site-specific cancers tested or overall cancer (**Fig. 2**). Two sets of genetic instruments were used to test all outcomes. The direction of the relationships in the FinnGen study was largely consistent with the direction of the associations in the UK biobank and the consortia (**Fig. S2 & S3**). Potential pleiotropic SNPs for site-specific cancers discovered by MR-PRESSO and leave-one-out analysis are displayed in **Table. S5**. In analyses using consortia data and UK biobank data, there was heterogeneity in the causal effects of SNPs for cancers including breast cancer, prostate cancer, non-melanoma skin cancer, pan cancer, colorectal cancer and lymphomas (Cochran’s-Q, P_het_ <0.05) (**Table. 1**); this was addressed after excluding rs6062486 and rs2581790 (associated previously with smoking) and outlying SNPs discovered by MR-PRESSO and leave-one-out analyses (**Table. 1**). After removing outliers, the results of association between AD and site-specific cancers were consistent with the original results using both Set 1-IVs and Set 2-IVs (**Table. 1**). In the FinnGen study, there was heterogeneity in causal effects across SNPs for non-melanoma skin cancer, this was addressed after excluding rs6062486 and rs2581790 (previously related with smoking) and outlying SNPs found by MR-PRESSO method and leave-one-out analyses (**Table. S6**). After excluding potential pleiotropic SNPs, the results remained consistent with the original results using both two sets of IVs (**Table. S6**). To further improve the power and strengthen our findings, we conducted meta-analysis of cancer sites with different data sources using both fixed and random effect models, we found the combined results largely reflected those for the consortia (where available) as these had many more cases than the two cohort studies. As in the single cohort/consortia analyses we did not find any strong evidence of effect of AD oncancer risk in any of our meta-analyses using both Set 1-IVs and Set 2-IVs (**Fig. 3 & Fig. S4**).

**Table 1.** Link between IVs for atopic dermatitis and risk of site-specific cancers, assessed in two ways.

| Outcome | Data Source | Full instruments (Set 1-IVs<sup>a</sup>)<br>OR (95% CI) | FDR | P for het<sup>b</sup> | Excluding pleiotropic SNP<br>OR (95% CI) | FDR | P for het<sup>b</sup> |
| --- | --- | --- | --- | --- | --- | --- | --- |
| Overall cancer | UK Biobank | 1.00 (0.94, 1.06) | 0.98 | P<0.001 | 1.03 (0.98, 1.08) | 0.57 | 0.20 |
| Breast cancer | BCAC | 0.97 (0.91, 1.03) | 0.80 | P<0.001 | 0.98 (0.95, 1.02) | 0.68 | 0.29 |
| Prostate cancer | PRACTICAL | 1.02 (0.92, 1.13) | 0.86 | P<0.001 | 0.99 (0.95, 1.04) | 0.85 | 0.50 |
| Ovarian cancer | OCAC | 0.94 (0.88, 1.00) | 0.51 | 0.38 | 0.95 (0.89, 1.02) | 0.57 | 0.69 |
| Non-melanoma skin cancer | UK Biobank | 1.04 (0.94, 1.16) | 0.80 | P<0.001 | 0.95 (0.85, 1.07) | 0.67 | 0.05 |
| Breast cancer | UK Biobank | 0.95 (0.87, 1.03) | 0.77 | 0.05 | 0.97 (0.88, 1.06) | 0.68 | 0.07 |
| Endometrial cancer | O'Mara et.al | 1.07 (0.98, 1.16) | 0.59 | 0.25 | 1.05 (0.97, 1.14) | 0.57 | 0.35 |
| Lung cancer | ILCCO | 1.00 (0.91, 1.10) | 0.99 | 0.25 | 1.00 (0.90, 1.11) | 0.97 | 0.19 |
| Prostate cancer | UK Biobank | 1.03 (0.91, 1.15) | 0.85 | P<0.001 | 0.96 (0.89, 1.04) | 0.67 | 0.71 |
| Colorectal cancer | UK Biobank | 0.90 (0.77, 1.07) | 0.80 | P<0.001 | 0.90 (0.79, 1.03) | 0.57 | 0.08 |
| Melanoma | UK Biobank | 0.96 (0.82, 1.12) | 0.80 | 0.04 | 1.04 (0.90, 1.20) | 0.79 | 0.23 |
| Lung cancer | UK Biobank | 1.04 (0.90, 1.20) | 0.80 | 0.27 | 1.07 (0.93, 1.23) | 0.67 | 0.42 |
| Colon | UK Biobank | 0.86 (0.72, 1.04) | 0.59 | 0.05 | 0.89 (0.74, 1.07) | 0.57 | 0.09 |
| Lymphomas | UK Biobank | 0.86 (0.62, 1.20) | 0.80 | P<0.001 | 0.87 (0.71, 1.07) | 0.57 | 0.24 |
| Bladder cancer | UK Biobank | 1.06 (0.88, 1.28) | 0.80 | 0.67 | 1.03 (0.84, 1.25) | 0.88 | 0.75 |
| Leukaemia | UK Biobank | 0.91 (0.74, 1.12) | 0.80 | 0.29 | 0.90 (0.72, 1.13) | 0.67 | 0.23 |
| Ovarian cancer | UK Biobank | 0.96 (0.79, 1.17) | 0.86 | 0.51 | 0.98 (0.80, 1.21) | 0.93 | 0.50 |
| Rectum | UK Biobank | 1.10 (0.81, 1.50) | 0.80 | 0.01 | 1.10 (0.85, 1.41) | 0.68 | 0.21 |
| Head and neck cancer | UK Biobank | 1.06 (0.86, 1.30) | 0.80 | 0.96 | 1.04 (0.84, 1.29) | 0.85 | 0.95 |
| Outcome | Data Source | Full instruments (Set 2-IVs<sup>c</sup>)<br>OR (95% CI) | FDR | P for het<sup>b</sup> | Excluding pleiotropic SNP<br>OR (95% CI) | FDR | P for het<sup>b</sup> |
| Overall cancer | UK Biobank | 1.00 (0.97, 1.02) | 0.92 | P<0.001 | 1.01 (0.99, 1.03) | 0.65 | 0.15 |
| Breast cancer | BCAC | 0.99 (0.96, 1.02) | 0.80 | P<0.001 | 0.99 (0.97, 1.02) | 0.79 | 0.39 |
| Prostate cancer | PRACTICAL | 1.01 (0.97, 1.06) | 0.80 | P<0.001 | 1.00 (0.97, 1.03) | 0.99 | 0.24 |
| Ovarian cancer | OCAC | 0.95 (0.91, 0.99) | 0.35 | 0.71 | 0.96 (0.92, 1.00) | 0.49 | 0.83 |
| Non-melanoma skin cancer | UK Biobank | 1.01 (0.97, 1.06) | 0.80 | P<0.001 | 1.01 (0.97, 1.04) | 0.85 | 0.40 |
| Breast cancer | UK Biobank | 0.98 (0.94, 1.02) | 0.80 | 0.36 | 0.98 (0.94, 1.02) | 0.67 | 0.28 |
| Endometrial cancer | O'Mara et.al | 1.02 (0.96, 1.08) | 0.80 | 0.07 | 1.01 (0.95, 1.07) | 0.85 | 0.10 |
| Lung cancer | ILCCO | 0.95 (0.90, 1.01) | 0.58 | 0.46 | 0.94 (0.89, 1.00) | 0.49 | 0.44 |
| Prostate cancer | UK Biobank | 1.00 (0.95, 1.06) | 0.97 | 0.01 | 0.95 (0.90, 1.00) | 0.49 | 0.76 |
| Colorectal cancer | UK Biobank | 0.92 (0.85, 0.99) | 0.35 | 0.01 | 0.94 (0.88, 1.01) | 0.52 | 0.17 |
| Melanoma | UK Biobank | 0.99 (0.92, 1.07) | 0.96 | 0.36 | 1.01 (0.94, 1.09) | 0.9 | 0.42 |
| Lung cancer | UK Biobank | 1.04 (0.96, 1.14) | 0.80 | 0.57 | 1.05 (0.97, 1.15) | 0.57 | 0.63 |
| Colon | UK Biobank | 0.88 (0.81, 0.97) | 0.29 | 0.32 | 0.90 (0.83, 0.99) | 0.49 | 0.52 |
| Lymphomas | UK Biobank | 0.93 (0.81, 1.06) | 0.80 | P<0.001 | 0.94 (0.84, 1.04) | 0.57 | 0.63 |
| Bladder cancer | UK Biobank | 1.12 (0.98, 1.28) | 0.59 | 0.19 | 1.10 (0.96, 1.27) | 0.57 | 0.17 |
| Leukaemia | UK Biobank | 1.05 (0.93, 1.19) | 0.80 | 0.40 | 1.14 (1.00, 1.30) | 0.49 | 0.82 |
| Ovarian cancer | UK Biobank | 0.94 (0.82, 1.07) | 0.80 | 0.37 | 0.94 (0.82, 1.08) | 0.67 | 0.33 |
| Rectum | UK Biobank | 1.02 (0.86, 1.20) | 0.93 | 0.01 | 1.05 (0.91, 1.21) | 0.68 | 0.31 |
| Head and neck cancer | UK Biobank | 0.96 (0.84, 1.10) | 0.80 | 0.79 | 0.95 (0.83, 1.09) | 0.68 | 0.74 |

Note (a). Set 1-IVs: Significant SNPs extracted from atopic dermatitis GWAS ( $P < 5E-08$ ; LD $r^2 < 0.001$ , 10000kb).
Note (b). P for het: P for heterogeneity
Note (c). Set 2-IVs: Significant SNPs extracted from atopic dermatitis GWAS ( $P < 5E-06$ ; LD $r^2 < 0.001$ , 10000kb).
BCAC, Breast Cancer Association Consortium; CI, confidence interval; ILCCO, International Lung Cancer Consortium; OCAC, Ovarian Cancer Association Consortium;
OR, odds ratio; PRACTICAL, Prostate Cancer Association Group to Investigate Cancer Associated Alterations in the Genome consortium.

**Figure 2.**
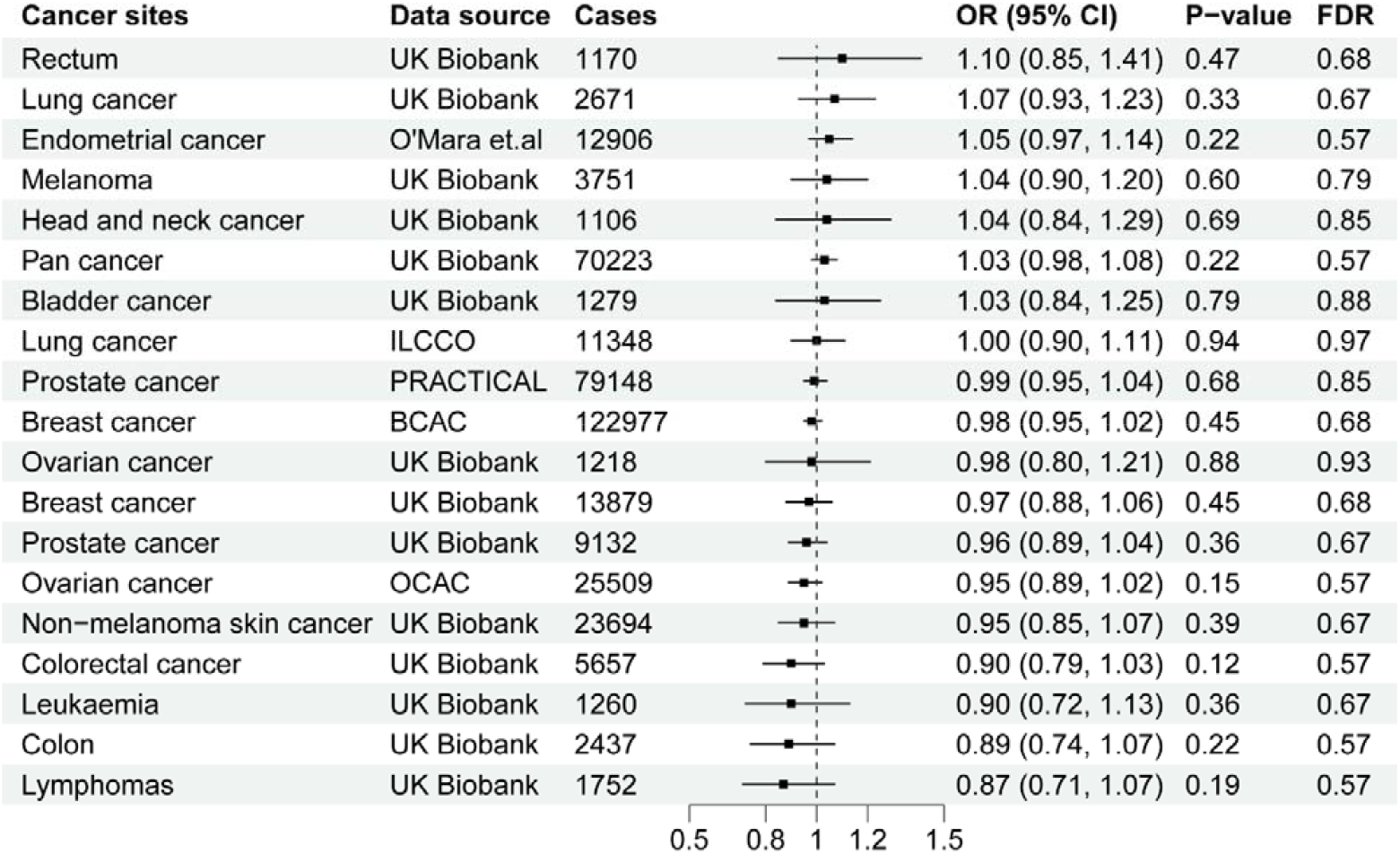
Associations of genetic predisposition to atopic dermatitis with site-specific cancers (Set 1-IVs). Set 1-IVs, IVs with the cutoff of P<10E-08. FDR, the adjusted P-value by the Benjamini-Hochberg method. BCAC, Breast Cancer Association Consortium; CI, confidence interval; ILCCO, International Lung Cancer Consortium; OCAC, Ovarian Cancer Association Consortium; OR, odds ratio; PRACTICAL, Prostate Cancer Association Group to Investigate Cancer Associated Alterations in the Genome consortium.

**Figure 3.**
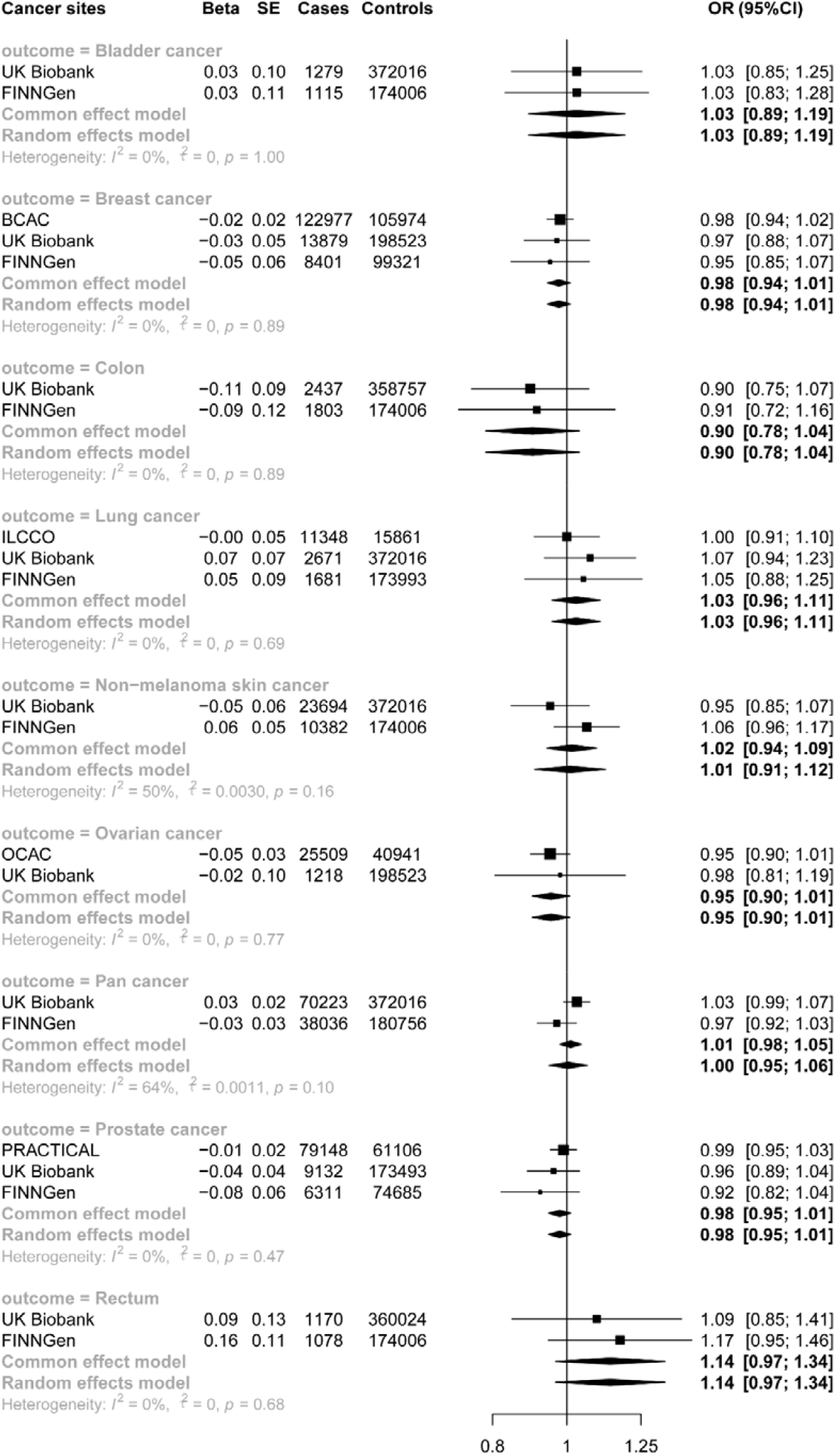
Meta-analysis of associations of genetic liability to AD and site-specific cancers with different sources of data using Set 1-IVs. BCAC, Breast Cancer Association Consortium; CI, confidence interval; ILCCO, International Lung Cancer Consortium; OCAC, Ovarian Cancer Association Consortium; OR, odds ratio; PRACTICAL, Prostate Cancer Association Group to Investigate Cancer Associated Alterations in the Genome consortium.

Results were broadly consistent in sensitivity analyses using alternative mendelian randomization methods, (**Table. S7 & S8**). In order to further assess the plausibility of confounding by violations of the independence assumption and the robustness of our results^29,30^, we ran analyses using positive and negative controls^20^, using both Set 1-IVs and Set 2-IVs (**Table. S9**). Consistent with current evidence of a positive link between AD and asthma^31^, we found a strong causal association between the genetically-predicted AD (Set 1-IVs) and asthma (OR, 1.34; 95%CI, 1.20-1.51; P<0.001). As expected, we observed no indication of a link between genetically predicted AD and our negative control outcome of body height (OR, 0.97; 95%CI, 0.90, 1.05; P=0.45). These results further strengthened our findings.

## Discussion

In this MR study on AD and risk of cancer, we systematically assessed the relationship of genetically predicted AD with a wide range of cancer outcomes. After applying the Benjamini-Hochberg correction, we found no indication of a link between genetic susceptibility to AD and site-specific cancers or overall cancer in the cancer consortia, UK biobank or FinnGen studies. Based on our current findings, it appears that our data do not provide sufficient support for the conclusions drawn from some prior observational studies linking AD to an increased risk of certain cancers. However, it is important to note that the low statistical power of some analysis may limit our ability to detect a significant association in certain site-specific cancers.

In a systematic review and meta-analysis including 32 studies (290,563 adults and 9,014 children) an inverse association between AD and brain cancer was found^7^ (pooled OR, 0.77; 95%CI, 0.71–0.83); but our study was unable to look at brain tumours due to the small number of cases in both FinnGen and UK Biobank and the lack of publicly available consortia data. The meta-analysis found no evidence of an association between AD and other cancers, which is consistent with our findings. Recently, in two large observational studies that were carried out in England and Denmark^8^, the associations between AD and overall cancer were HR,1.04; 99% (CI, 1.02-1.06) in England and 1.05 (99% CI, 0.95-1.16) in Denmark suggesting a small increased risk with AD across all cancer sites, our analysis in the FinnGen cohort did show weak evidence of a small positive effect (OR,1.03; CI, 0.99-1.07) however our results for UK Biobank (OR, 0.97; CI, 0.92-1.03) and our meta-analysis results (OR,1.00; 95%CI,1.00-1.01) did not support this finding. In the same observational study,^8^ AD was linked to an increased likelihood of developing lymphoma in England. In another systematic review and meta-analysis including 8 population-based cohort studies (n = 5, 726,692) and 48 case-control studies (n = 14 136), Wang et al^6^. reported significant associations of AD and several site-specific cancers including keratinocyte carcinoma (pooled SIR, 1.46; 95% CI, 1.20-1.77), kidney cancer (pooled SIR, 1.86; 95% CI, 1.14-3.04), as well as cancer of lung and respiratory system (pooled OR, 0.61; 95% CI, 0.45-0.82). Zhu et al. conducted a systematic review consisting of 16 studies with a total of 9,638,093 participants in order to investigate the contribution of AD to skin cancers, this study showed that AD was associated with an elevated odds of nonmelanoma skin cancer but not associated with melanoma^32^. A previous MR study found no association between allergic diseases (composed of asthma, hay fever, and eczema), and the risk of prostate cancer and breast cancer, which aligns with our findings to a certain extent^33^. Based on our results, a causal connection between AD and the risk of cancers cannot be established. One explanation for this could be that the previously observed associations between AD and site-specific cancerswere chance findings or that they were confounded by an unknown factor. Alternatively, we may have failed to find strong evidence of associations due to low power in some of our analyses, because of this we were unable to rule out effects of AD on lymphoma and nonmelanoma skin cancer seen in the observational analyses and although our results for lung cancer were not consistent with the large protective effect seen in the meta-analysis by Wang et al^5^ we did see some evidence of a protective effect of AD on lung cancer using our set 2-IV. Due to small numbers of cases, we did not perform analyses of brain cancers and keratinocyte carcinomas which have been found to be associated with AD in observational studies.

Our study has several strengths. Firstly, this is the first report that systematically investigated the potential relationship between AD and a broad range of cancer sites using a MR design. Secondly, two-sample MR enabled us to use the largest GWAS of AD, and our study design is based on a discovery set and a validation set (both with two sets of IVs), which could promote the reliability of our findings. Thirdly, we performed a positive and negative control outcome MR analysis, both of which served to further confirm the appropriateness of the IVs used. One limitation of the current study lies in genetic summary statistics, which restricts the types of analyses of analysis that can be performed. In addition, statistical power was limited in some analyses of cancer cases. Another point is that our findings should be considered in the context of the fact that the AD status in our study is defined by genetic liability based on genetic variants, which might not be comparable to other previous observational studies. However, based on our observed and consistent null results from several complementary methods and data sources, it is less likely that our findings are distorted by bias.

In summary, we found no evidence of causal relationship for AD and overall cancer risk or any site-specific cancers. The possibility that treatment for AD could be involved in the chance of developing cancer should be the primary focus of future research.

## Data Availability

All data produced in the present work are contained in the manuscript

https://gwas.mrcieu.ac.uk/

## Acknowledgments

The authors would like to thank the participants of the individual studies contributing to the FinnGen, UK Biobank, Breast Cancer Association Consortium, the Ovarian Cancer Association Consortium, International Lung Cancer Consortium, and the Prostate Cancer Association Group to Investigate Cancer Associated Alterations in the Genome consortium.

## Declarations

### Ethics approval and consent to participate

The study followed the Strengthening the Reporting of Observational Studies in Epidemiology using Mendelian Randomization.

### Consent for publication

All authors read and approved the final manuscript for publication.

### Availability of data and materials

All supporting data are included in the manuscript and supplemental files. Additional data are available upon reasonable request to the corresponding author.

### Author contributions

Conception and design: Qiang Liu, Sarah J Lewis.

Collection and assembly of data: Qiang Liu, Yipeng Wang

Data analysis and interpretation: Qiang Liu

Manuscript writing, review and editing: Qiang Liu, Li Chen, Sarah J Lewis, Jing Wang

Final approval of manuscript: Qiang Liu, Li Chen, Yipeng Wang, Xiangyu Wang, Sarah J Lewis, Jing Wang

